# Impact of pathogen reduction methods on immunological properties of the COVID-19 convalescent plasma

**DOI:** 10.1101/2020.10.02.20205567

**Authors:** Alexander I. Kostin, Maria N. Lundgren, Andrey Y. Bulanov, Elena A. Ladygina, Karina S. Chirkova, Alexander L. Gintsburg, Denis Y. Logunov, Inna V. Dolzhikova, Dmitry V. Shcheblyakov, Natalia V. Borovkova, Mikhail A. Godkov, Alexey I. Bazhenov, Valeriy V. Shustov, Alina S. Bogdanova, Alina R. Kamalova, Vladimir V. Ganchin, Eugene A. Dombrovskiy, Stanislav E. Volkov, Nataliya E. Drozdova, Sergey S. Petrikov

## Abstract

**Background and Objectives:** COVID-19 convalescent plasma is an experimental treatment against SARS-CoV-2. The aim of this study is to assess the impact of different pathogen reduction methods on the levels and virus neutralizing activity of the specific antibodies against SARS-CoV2 in convalescent plasma.

**Materials and Methods:** A total of 140 plasma doses collected by plasmapheresis from COVID-19 convalescent donors were subjected to pathogen reduction by three methods: methylene blue (M)/visible light, riboflavin (R)/UVB, and amotosalen (A)/UVA. To conduct a paired comparison, individual plasma doses were divided into 2 samples that were subjected to one of these methods. The titres of SARS-CoV2 neutralizing antibodies (NtAbs) and levels of specific immunoglobulins to RBD, S- and N- proteins of SARS-CoV-2 were measured before and after pathogen reduction.

**Results:** The methods reduced NtAbs titres differently: among units with the initial titre 80 or above, 81% of units remained unchanged and 19% decreased by one step after methylene blue; 60% were unchanged and 40% decreased by one step after amotosalen; after riboflavin 43% were unchanged and 50% (7% respectively) had a one-step (two-step respectively) decrease. Paired two-sample comparisons (M vs A, M vs R and A vs R) revealed that the largest statistically significant decrease in quantity and activity of the specific antibodies resulted from the riboflavin treatment.

**Conclusion:** Pathogen reduction with methylene blue or with amotosalen provides the greater likelihood of preserving the immunological properties of the COVID-19 convalescent plasma compared to riboflavin.

## Introduction

The new coronavirus infection (COVID-19) caused by the SARS-CoV-2 virus continues its march around the world, causing a global crisis as the number of new cases and deaths continues to rise. The treatment is supportive care mostly aimed at relieving symptoms. Candidate vaccines are still going through different stages of clinical trials, and different classes of drugs are being tested to inhibit virus replication and reduce inflammation [1, 2, 3].

Passive immunotherapy with polyclonal antibodies from the blood plasma of convalescents was tested earlier in the outbreaks of SARS-CoV, influenza and other dangerous infections of the twentieth century [4, 5, 6]. Experts from different countries came to consider the possibility of using COVID-19 convalescent plasma (CCP) for therapeutic purposes in patients with COVID-19 [7, 8, 9]. In many countries, national campaigns have been launched to collect CCP. The use of CCP is still exploratory at this time as proof of efficacy has not been demonstrated through RCT even if many RCTs on CCP are ongoing worldwide.

Various mechanisms have been suggested as responsible for the therapeutic effect of CCP such as virus neutralization and immunomodulation [10]. Virus neutralizing antibodies (NAbs) of IgG, IgM and IgA classes bind to different parts of glycoprotein S, including the region of the receptor-binding domain (RBD), spatially blocking its interaction with the membrane protein ACE2 of host cells, which limits the penetration of the virus into the cell, thereby limiting viral replication [11, 12, 13]. Along with antibodies against different fragments of S-protein antibodies against nucleocapsid (N-protein) are detected in the course of COVID-19 infection. These antibodies are used as additional diagnostic markers, but do not correlate with virus neutralizing activity *in virto* [14].

Every plasma transfusion is associated, however, with risks of virus transferral such as HIV, HBV, HCV, etc. [15]. Donor studies of Cappy et al [16] showed that viremia was extremely rare in asymptomatic blood donors, viral RNA levels were very low when detected, and the corresponding plasma was not infectious in cell culture. At the moment, there are no scientific publications reporting on the transmission of SARS-CoV- 2 through the transfusion of blood components [17]. The Working Party on Global Blood Safety of the International Society of Blood Transfusion (ISBT) recommended the use of pathogen reduction (PR) of convalescent plasma to minimize the residual risk of blood-borne infections and to address the problem of possible superinfection with the SARS-CoV-2 virus [8].

Until recently, there was surprisingly very little information on the effect of PR treatment of plasma on the functional properties of immunoglobulins. This issue has been raised previously for Ebola convalescent plasma regarding possible impact of PR by Intercept technology on the neutralizing activity of EBOV IgG, potentially affecting clinical outcomes [18, 19]. Tonn *et al* [20] found that PR did not impair the stability and neutralizing capacity of SARS-CoV-2 specific antibodies in 5 CCP units treated with psoralen/UVA (Intercept).

To date, there is no sufficient data on how pathogen reduction affects the immunological properties of CCP and what PR technologies are preferable to use to maintain its quality and effectiveness.

The objective of this study is to assess the effect of various methods for pathogen reduction on the levels and virus neutralizing activity of the specific antibodies against SARS-CoV2 in CCP.

## Materials and methods

The COVID-19 convalescent plasma procurement program in Russia was launched on April 2, 2020 at the Department of Transfusion Medicine of the Sklifosovsky Research Institute of Emergency Medicine, Moscow. At present, this program involves many hospitals in several regions and has more than 6,500 donations and about 4,500 transfusions of CCP in Moscow alone. According to the adopted regulations, donors of convalescent plasma were recruited among the individuals with prior diagnosis of COVID-19 infection documented by a positive RT-PCR-test who received treatment either in a hospital setting or on an outpatient basis. Donors fulfilled the standard blood donor selection criteria. Plasma was collected at least 2 weeks after the complete disappearance of clinical symptoms.

Ethical approval was granted for this study by the Independent Moscow City Research Ethical Committee in accordance with national regulations. Informed consent was obtained in writing from all donors prior to donation.

Plasmapheresis procedures were performed using Auto-C (Fresenius Kabi), Aurora (Fresenius Kabi) and PCS2 (Haemonetics) machines. Plasmapheresis was carried out in accordance with standard protocols collecting an amount of 650 ml plasma. During collection, the same anticoagulant (ACD-A) was added in a ratio of 1:12 in all machines.

Pathogen reduction procedures were carried out immediately after the end of plasmapheresis. For comparison, three systems for PR were selected:

1. Intercept (Cerus): 15 mL of 6 mmol/L amotosalen hydrochloride solution were mixed with plasma to a final concentration 150 µmol/L and exposed to UVA light (3 J/cm^2^), thereafter residual amotosalen and free photoproducts were adsorbed in a compound adsorption device (CAD).
2. Mirasol (Therumo, BCT): 35 mL of 500 µmol/L riboflavin solution was mixed with 200±5 mL plasma and exposed to UVB light (6.24 J/mL).
3. Maco-Tronic (MacoPharma): plasma was transferred to the THERAFLEX MB-Plasma illumination bag, passing through a chamber containing the anhydrous MB pill, resulting in a minimum MB concentration of at least 0.8 µmol/L in 315 mL of plasma, subsequent illumination (120 J/cm^2^) and removal of MB using the Blueflex MB removal filter.

All three PR technologies were validated at our blood bank for routine use, prior to pathogen reduction of CCP for this study. All plasma units were treated under the specific PR manufacturer’s instructions and met the required specifications for each pathogen reduction technology. The temperature of plasma units during illumination were maintained at ≤22C.

After PR plasma was frozen at -40□C and became available for clinical use after receiving negative results of all serology/virology tests for transfusion-transmitted diseases.

The study included 140 doses of plasma obtained by plasmapheresis from 140 COVID- 19 convalescent donors. From each plasma unit, samples were collected before and after pathogen reduction for the determination of the titres of SARS-CoV-2 neutralizing antibodies (NtAbs), as well as quantitative determination of specific IgG to the receptor-binding domain (RBD) of the glycoprotein S of the SARS-CoV-2 virus, and specific IgM and IgG to S- and N- proteins of this virus.

To conduct a paired two-sample comparison in order to assess the effect of each of these methods on the immunological parameters of CCP, the plasma dose from each donor was divided into 2 parts, and each part was then simultaneously subjected to a pathogen reduction procedure by one of the two methods according to the following scheme:

- pair 1: methylene blue (M) vs. riboflavin - 48 pairs;
- pair 2: amotosalen (A) vs. riboflavin (R) - 36 pairs;
- pair 3: methylene blue (M) vs. amotosalen (A) - 56 pairs;

Since there were 140 samples *before* treatments, in total 420 (=48*2+36*2+56*2+140) samples were analysed. Virus neutralization assay and ELISA were performed at a day of biomaterial collection at the Gamaleya National Research Center of Epidemiology and Microbiology, Moscow. Samples for chemiluminescent immunoassay (CLIA) were frozen immediately after the collection in Eppendorf tubes in aliquots of 200 µL in -35□C and thawed and analysed later all at the same time.

NtAb titer was determined by microneutralization test [21]. Vero E6 were cultured in DMEM - Dulbecco’s Modified Eagle Medium (Gibco) containing 10% heat-inactivated fetal bovine serum (Gibco), 3,7 g/l sodium bicarbonate (PanEco), 1 mM glutamine (PanEco), 100 µg/ml streptomycin (PanEco), 100 IU/ml penicillin (PanEco) in 5% CO_2_ humidified incubator at 37°C.

The SARS-CoV-2 (hCoV-19/Russia/Moscow_PMVL-1/2020) virus was obtained from the State Virus Collection of Gamaleya NRCEM. The infectious virus titre was determined on Vero E6 cells using a 50% tissue culture infectious dose (TCID50) assay. Serial 10-fold dilutions of the virus stock were prepared in DMEM with 2% heat-inactivated FBS and in volume of 100 µl were added to Vero E6 cells in a 96-well plate in 8 repeats. The cells were incubated at 37°C in 5% CO_2_ for 96 hours and scored visually for cytopathic effect. The TCID50 titre was calculated by Reed and Muench method. Neutralization activity of plasma was determined by microneutralization test using SARS-CoV-2 virus in a 96-well plate. Plasma samples were inactivated by incubation at 56°C for 30 min and serial two-fold dilutions in DMEM containing 2% heat-inactivated FBS at a range 1:20 – 1:1280 were made. Then 100 TCID_50_ was added to each sample. The samples were incubated at 37°C for 1 h in a 5% CO2 incubator. After incubation samples were added to Vero E6 cells and incubated in a 5% CO2 incubator at 37°C for 96 h. Neutralization titre was defined as the highest serum/plasma dilution without cytopathic effect in two of three replicable wells.

Anti-SARS-CoV-2 IgG semi-quantitative ELISA test-system developed in Gamaleya NRCEM and registered for clinical use in Russian Federation (P3H 2020/10393 2020-05-18) was used for the determination of the IgG specific to the receptor-binding domain of SARS-CoV-2 glycoprotein S [20].

Briefly, the RBD-pre-coated plates (100 ng per well) were washed 5x with 0.1% wash solution and then blocked with blocking solution. Plasma samples were diluted 1/200 in blocking solution and added in wells, then plates were incubated at 37°C for 1 h. After washing the plates 5x the peroxidase-conjugated anti-human IgG detection antibodies diluted in blocking solution were added and plates were incubated at 37°C for 1 h. After washing 5x the substrate TMB was added and plates were incubated at 20- 25°C for 15 min, the reaction was stopped with the stop solution. The OD signals were determined with a spectrophotometer Multiskan FC (Thermo Fisher Scientific Inc, USA) at 450 nm.

Sensitivity of the test is 96%, specificity 100% and the limit of detection is 0.2 AU (OD450nm).

The CL-series SARS-CoV-2 IgG and IgM assays are a two-step chemiluminescent immunoassays for detection of IgG and IgM SARS-CoV-2 antibodies in human serum or plasma, performed on the fully automated Mindray CL 1200i analytical system (Shenzhen Mindray Bio-Medical Electronics Co., Shenzen, China). Samples react with paramagnetic microparticles coated with SARS-CoV-2 specific antigens - recombinant N-Protein and Spike (S) Protein. Alkaline phosphatase-labeled anti-human IgG or IgM monoclonal antibodies are added to the reaction to form a sandwich with microparticles captured anti-SARS-CoV-2 antibodies. Finally, a substrate solution is added, resulting in a chemiluminescent reaction measured as relative light units by a photomultiplier built into the system. The amount of SARS-CoV-2 IgG antibodies present in the sample is proportional to the relative light units (RLUs) generated during the reaction. The SARS-CoV-2 IgG and IgM antibodies concentration can be determined via a calibration curve, which is built on an encoded Master Calibration Curve and three level product calibrators. Cut-off values are: IgG positive□>□10.0 U/mL and IgM positive > 1.0 COI, Positive Percent Agreement 81.7% and 82.2%, Negative Percent Agreement 91.6% and 94.9%for IgM and IgG respectively, according to the manufacturer’s specifications.

Since the data for NtAbs are presented in the format 10 multiplied by an integer power of two (i.e., 20, 40, 80, 160 etc.) we log-2-transformed the data: y=log2(x/10), where x is the reported value of NtAbs

To identify the methods of pathogen reduction which have the least negative effect on NtAbs levels, we applied the two-sample paired T-tests (M vs A, M vs R, and A vs R) to the difference in reduction in titres of NtAbs, anti-RBD IgG, and anti-S + N IgG and IgM titres respectively, after pathogen reduction by different methods. The p-values < 0.05 were considered to be significant. No corrections were made for multi-significance.

Confidence intervals were obtained using the standard methods for estimation of proportions. The software used for the analysis was Maple™.

## Results

The assessment of the impact of various methods of pathogen reduction on the titres of SARS-CoV2 neutralizing antibodies (NtAbs) showed a statistically significant decrease in antibody titres after all pathogen reduction processes (Figure 1, Table 1).

**Table 1:**
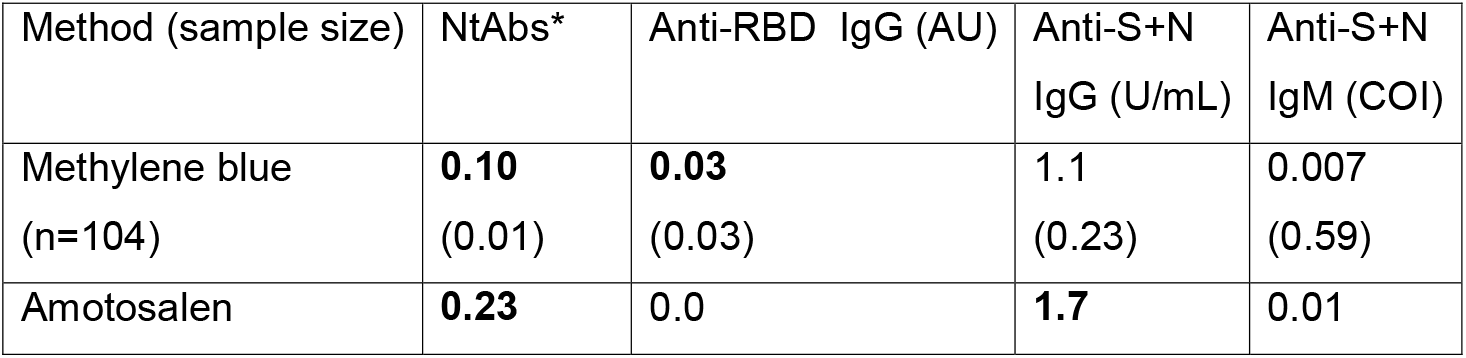
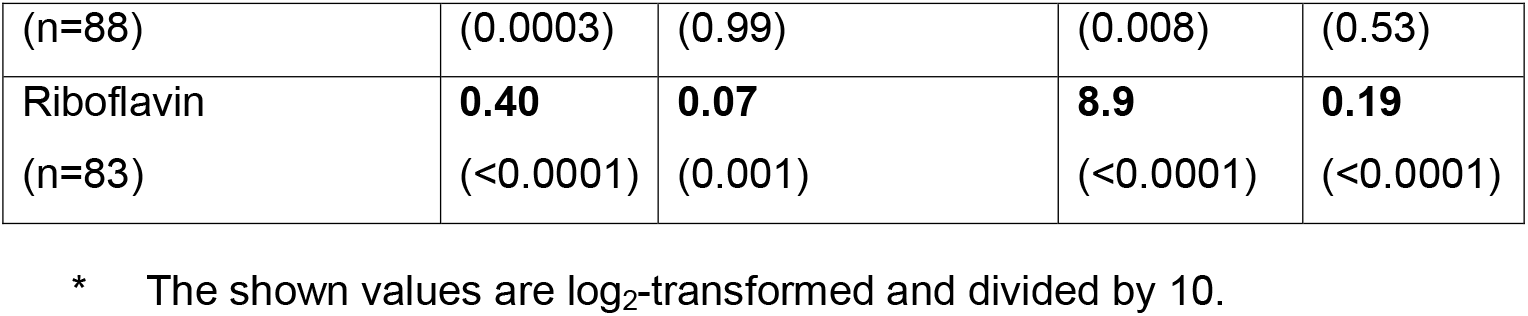
Individual methods of pathogen reduction, comparison of the initial values *vs*. post treatment values using two sample paired t-test. The numbers show the average decline of values of NtAbs, Anti-RBD IgG, Anti S+N IgG and Anti S+N IgM after treatments by each of the three methods (M/A/R respectively). The p-values in the parentheses below indicate how statistically different from zero these values are; those with p<0.05 are in bold font.

**Figure 1.**
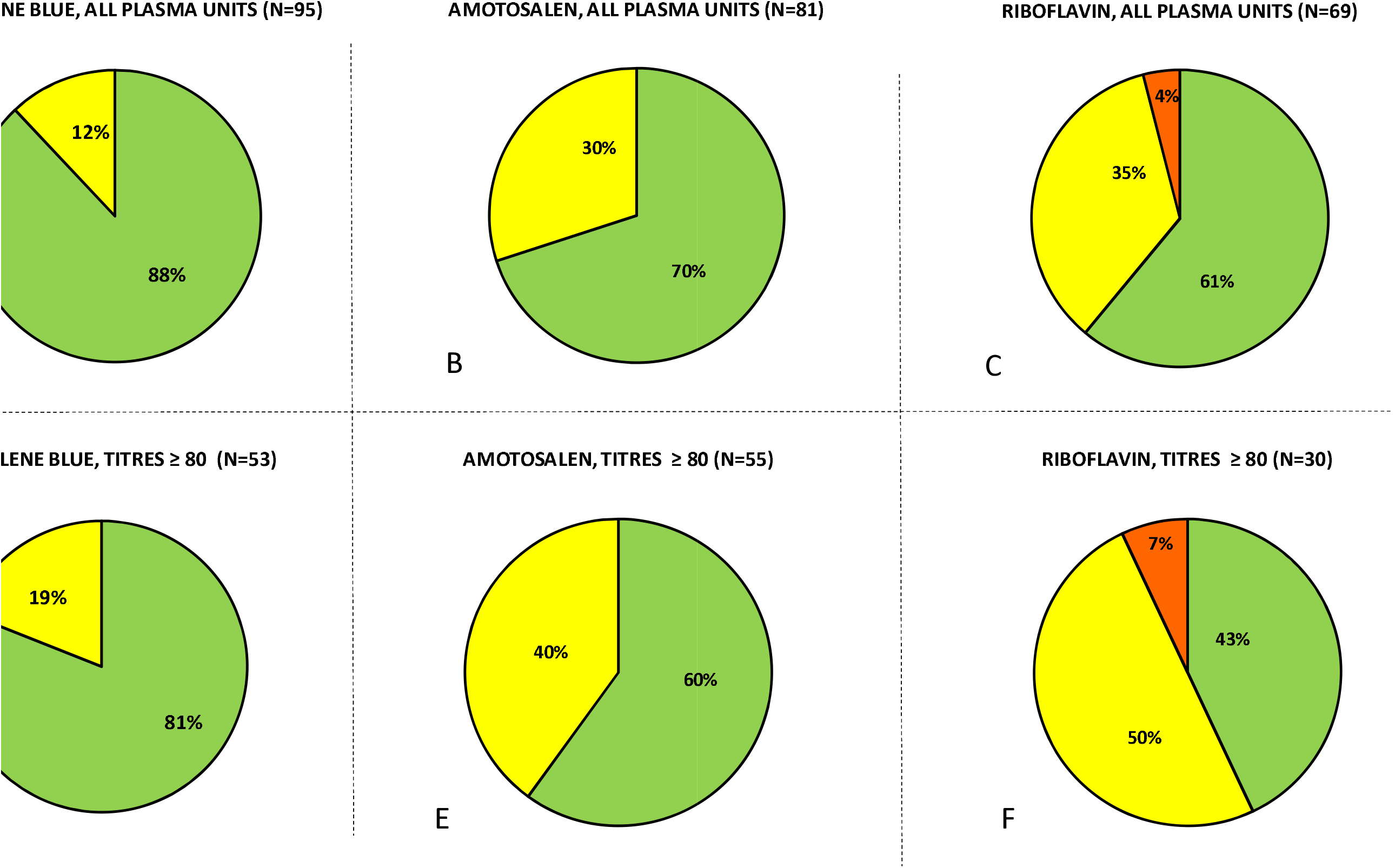
Percentage of COVID-19 convalescent plasma units that had no decrease in NtAbs titre 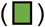, one-step titre decrease 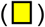 or two steps titre decrease 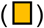 after pathogen reduction with methylene blue (A, D), amotosalen (B, E) and riboflavin (C, F) among all plasma units (A, B, C) or only units with initial NtAbs titre 80 or above (D, E, F).

If all plasma units, regardless of the initial titre were included in the analysis, it was shown that in 88% (n=104; confidence interval 81% - 94%) of units NtAbs titres did not decrease after pathogen reduction with methylene blue whereas a one-step titre reduction was observed in remaining 12%. In 70% (n=88; 95% confidence interval 61% - 80%) of units treated with amotosalen the NtAbs titre did not change, and in 30% it decreased by 1 step. Pathogen reduction with riboflavin left NtAbs titres unchanged in 61% (n=83; confidence interval 51% - 72%) of the units, in 35% decreased by one step and in 4% by two steps.

To compare the impact of different methods of pathogen reduction, we used the data collected on paired data: the plasma units from the same donor were treated using e.g. method A and M, and then the resulting NtAbs were noted for both methods. We had three different datasets: one compared A vs. R, another dataset M vs. R and the third M vs. A. The results are the following: M is better than R (p-value=0.00002, n=48), A is better than R (p-value=0.0002, n=36), M is better than A (p-value=0.0012, n=56).

When only units with the initial NtAbs titre 80 or above were chosen (the level considered to be suitable for therapeutic purposes) the distribution was similar: after treatment with methylene blue, 81% of plasma samples had unchanged NtAbs titres (n=53; confidence interval 71% - 92%), while in the remaining 19% of samples the titres decreased by 1 step. Pathogen reduction with amotosalen gave worse results: 60% of samples had the same NtAbs after the reduction (n=55; confidence interval 47% - 73%), while in the remaining 40% samples the titres decreased by 1 step. Finally, after treatment with riboflavin, only 43% of the samples preserved the level of NtAbs titres (n=30; confidence interval 26% - 61%), whereas a one-step decrease was observed in 50% samples, and a two-step decrease in 7% of samples.

The decrease in anti-RBD IgG in paired comparison with baseline values was most pronounced after pathogen reduction with riboflavin followed by methylene blue whereas after amotosalen there was no significant difference (Table 1).

Plasma pathogen inactivation with methylene blue did not lead to a significant decrease in anti-S + N IgG and IgM, whereas the use of amotosalen significantly reduced only the level of anti-S + N IgG (Table 1). In the study of 83 pairs of samples before and after pathogen reduction with riboflavin, the differences were significant in the anti-S + N levels of both IgG and IgM (Table 1).

Paired two-sample comparisons (M vs A, M vs R and A vs R) revealed the most prominent and statistically significant decline in titres of NtAbs, anti-S + N IgG and IgM (with the exception of anti-RBD IgG titres) resulted from pathogen reduction by riboflavin compared with two other PR technologies (Table 2).

**Table 2:** Comparison of different methods of pathogen reduction using two sample paired t-test. The numbers show the average differences of declines of values of NtAbs, Anti-RBD IgG, Anti S+N IgG and Anti S+N IgM corresponding to the respective pair of methods. The p-values in parentheses below indicate how statistically different from zero these values are; those with p<0.05 are in bold font.

| Method<br>(sample size) | NtAbs* | Anti-RBD IgG<br>(AU) | Anti S+N IgG<br>(U/mL) | Anti-S+N<br>IgM<br>(COI) |
| --- | --- | --- | --- | --- |
| Methylene blue vs<br>Amotosalen (n=56) | **0.27**<br>(0.001) | -0.02<br>(0.60) | 3.1<br>(0.21) | 0.045<br>(0.29) |
| Amotosalen vs Riboflavin<br>(n=36) | **0.33**<br>(0.0002) | 0.06**<br>(0.27) | **6.7**<br>(<0.0001) | **0.20**<br>(<0.0001) |
| Methylene blue vs<br>Riboflavin (n=48) | **0.42**<br>(<0.0001) | 0.007<br>(0.82) | **6.6**<br>(<0.0001) | **0.16**<br>(<0.0001) |

\* The shown values are log<sub>2</sub>-transformed and divided by 10.
\*\* one data point is missing, so n=35 here

As the riboflavin is not removed after the illumination phase in the Mirasol technique, the residual amount of riboflavin or its by-products may possibly affect the *in vitro* assessment of the NtAbs. To rule out this possibility we have conducted a series of tests showing that the viability of the cells used to assess the neutralization was not affected by riboflavin itself nor its derivatives. We also measured NtAbs titres in several plasma samples taken after the addition of riboflavin but before the illumination - the titres did not differ from those in samples before PR. Thus, it seems that the reducing effect of PR with riboflavin on the level and activity of antibodies against SARS-CoV-2 requires both photosensitizing agent and UVB illumination as this PR technology requires.

## Discussion

The key safety issue of using convalescent plasma is played by the choice of a PR technology that minimizes the residual transfusion risk of transmissible viruses in the final product, while maintaining a high titre of antibodies to the SARS-Cov-2 virus. A number of different PR technologies are available today [22]. Ultraviolet (UV) A [23, 24] and UVB radiation [25], in combination with amotosalen and riboflavin, respectively, makes it possible to inactivate nucleic acids of pathogenic organisms. These systems can reduce the activity of SARS and MERS viruses in plasma or platelet concentrates to varying degrees. Methylene blue is a phenothiazine compound that, in combination with visible light, is also capable of inactivating coronaviruses in plasma [26, 27]. The photoactive agents used in these methods have different chemical structures and are activated at different wavelengths of radiation (visible light with the peak wavelength of 590 nm, UVA from 400 nm - 315 nm and UVB from 315 nm - 280 nm). Consequently, various mechanisms are involved in ensuring pathogen reduction. The amotosalen intercalates into DNA and RNA and, when activated by UVA light, causes covalent cross-linking of those nucleic acids thus preventing the replication. The riboflavin binds to nucleic acids and, when activated by the illumination step, alters guanine residues via type I and type II redox reactions. MB can intercalate into DNA or bind to the DNA helix, depending on the concentration and ionic strength of Mg^2+^. When exposed to light type I (redox) or type II (photo-oxidative) reactions occur, with most of the PR activity resulting from type II reactions [22, 28].

In earlier studies on Ebola convalescent plasma [18, 19] it was shown that PR with amotosalen/UVA only slightly reduced anti-Ebola virus IgG titres and Ebola-specific neutralizing antibodies. Tonn *et al* [20] found that PR did not impair the stability and neutralizing capacity of SARS-CoV-2 specific antibodies in 5 CCP units treated with psoralen/UVA.

The current study is the first to compare the impact of different PR technologies on SARS-CoV-2 antibody levels and activity in convalescent plasma.

The hypothesis tested in this study is that different types of photo-chemical reactions used in standard PR technologies can have a different effect on the amount and neutralizing activity of SARS-CoV2 specific antibodies in the final product - convalescent plasma. The results obtained indicate a lesser effect on the immunological quality of CCP of pathogen reduction with methylene blue or with amotosalen, possibly due to the lesser amount of energy used for illumination and lesser amount of reactive oxygen species releasing after photoactivation compared with riboflavin [28, 29]. More research is needed to elucidate the exact mechanisms for the oxidative damage of proteins and particularly immunoglobulins in course of different PR technologies.

Based on the study, we can recommend using pathogen reduction with methylene blue or with amotosalen to ensure the safety and quality of CCP, due to the greater likelihood of preserving the immunological properties of the final product. Since even these technologies are associated with a risk of reducing the quantity and quality of antibodies against SARS-CoV-2, it is recommended to transfuse at least 2 units of convalescent plasma (200-300 ml) from different donors to one patient, especially in those medical institutions where the routine measurement of NtAbs titres is not possible.

In those blood establishments where pathogen reduction with riboflavin is traditionally used, it may be worth to consider increasing the dose of transfused convalescent plasma in order to compensate for the decrease in the baseline neutralizing antibody titres after this method of pathogen reduction.

## Data Availability

All the data referred to at this manuscript is available

## Acknowledgements

The authors thank the medical and laboratory staff of the Department of Transfusion Medicine of the Sklifosovsky Research Institute of Emergency Medicine and at the Gamaleya National Research Center of Epidemiology and Microbiology without whose assistance the study would not have been possible. They also thank Senior Consulting Physician Jens Kjeldsen-Kragh, Department of Clinical Immunology and Transfusion Medicine, Region Skane Medical Service, Lund, Sweden and also two anonymous reviewers from Vox Sanguinis for carefully reading this paper and making many valuable suggestions.

## Conflict of interest

The authors declare no conflict of interests.

## Notes

### Competing Interest Statement

The authors have declared no competing interest.

### Funding Statement

This study was made possible in the context of Moscow Government COVID-19 Convalescent Plasma Program. Stanislav Volkovs research is partially supported by the Swedish Research Council and the Crafoord foundation.

### Author Declarations

Ethical approval was granted for this study by the Independent Moscow City Research Ethical Committee in accordance with national regulations.

### Summary of Updates

Abstract was rewritten for clarity, severan important references were added in the Introduction and i the Discusson, more detailed description of all the methods used were given in Materials and Methods as well as information concerning ethical approval and donor consent. All the results values are unchanged. Some new aspects added to the Discussion and several references are added to References.

